# TAPSE, Tissue Doppler Systolic annular velocity(S’) and myocardial performance Index (MPI) as a predictor of proximal RCA occlusion in acute inferior wall myocardial ST elevation infarction

**DOI:** 10.1101/2023.08.14.23294098

**Authors:** Sanjeev Kumar, Chandra Bhan Meena, Navida Babbar, Himanshu Mahla, Shashi Mohan Sharma, Rajeev Bagarhatta, V.V. Agrawal, Vijay Pathak, Deepak Maheshwari, Dhananjay Shekhawat, Ankita

**Author notes:** Corresponding author-, Address – Plot no -613, Gadhwalo ki Dhani, Chhawashri, Gudha Gauji, Jhunjhunu -333012 Rajasthan, India. Department of Dermatology, SMS Medical College Jaipur, Rajasthan, 302004 India.

## Abstract

**Objectives:**

- Assessment of RV function to predict proximal RCA stenosis and identification of higher-risk patients in IWMI by echocardiographic parameters.

**Study design:** Two-arm, single-center cross-sectional study in SMS medical college and Hospital Jaipur.

**Methods:** We enrolled 67 patients with first episode of acute IWMI within 24 hours of symptom onset. TAPSE, S’, and MPI were measured. Within one month, patients underwent coronary angiogram or intervention. Based on angiographic findings patients divided into group A (significant proximal RCA stenosis) and group B (without significant proximal RCA stenosis).

**Results:** 25 patients were in group A and 42 in group B. RV involvement was significant among patients with proximal RCA stenosis. A significantly low SBP, DBP, HR, and raised JVP, NT-pro BNP, Troponin I with ST elevation V4R, RV diastolic dysfunction, arrhythmia, and underwent intervention were found among the group with proximal RCA stenosis.

Patients with proximal RCA stenosis had significantly low TAPSE-M, ET-PW, ET by TDI, S’, E’, RVFAC, and high MPI by PW, IRT by TDI, ICT by TDI, MPI by TDI on ECHO.

Sensitivity to detect proximal RCA stenosis by TAPSE-M mode 92%, MPI by TDI 88%, and 100% specificity by S’ by TDI < 10 cm/s, MPI by TDI, and ST elevation V4R.

**Conclusion:** Echocardiographic assessment of RV parameters like TAPSE, S’ velocity, and MPI are useful in predicting proximal RCA as infarct-related artery in IWMI. Patients with proximal RCA stenosis had raised JVP, NT-proBNP, Troponin I, ST elevation V4R, RV diastolic dysfunction, arrhythmia and underwent intervention.

## Introduction

- RV infarction is one of the major cause of RV contractile dysfunction. RV infarction occurs in 20–50% of inferior myocardial infarctions. [1] Patients with right ventricular infarctions associated with inferior myocardial infarctions have much higher rates of bradycardia requiring pacing support, significant hypotension, and in-hospital mortality than isolated inferior myocardial infarction. [2]
- Occlusion of the proximal dominant right coronary artery is usually responsible for right ventricular infarction in inferior wall myocardial infarction. [3] The classic clinical triad of right ventricular infarction includes distended neck veins, clear lung fields, and hypotension. Electrocardiogram is often inadequate to predict proximal right coronary artery stenosis as infarct-related artery. [4]
- Electrocardiogram changes are transient and disappear in 48% of cases within 10 h, making it a less dependable tool. [5] Echocardiography, being non-invasive, comparatively inexpensive, widely available, and having no side effects is the modality of choice for the assessment of morphology and function of the RV in clinical practice. [6]
- Conventional measurements of area and volume have limited utility in assessing RV function[7] due to the complex geometry of the right ventricle and difficulty in defining the endocardial borders. [8]
- Using Doppler myocardial imaging, several regional and global parameters such as direction, timing, and amplitude of the velocity of the ventricular wall can be determined. This technique is less dependent on chamber geometry. Furthermore, endocardial border delineation is not required, which makes TDI more useful even if the echocardiographic image quality is suboptimal. [6]

### Aims & Objectives

- Assessment of RV function to predict proximal RCA stenosis by Echocardiographic parameters.
- Identify a subset of patients with inferior wall myocardial infarction at higher risk of unfavorable clinical events.

## Material and method

- Our study was a hospital-based, two-arm, single-center cross-sectional study.
- This study was done from March 2022 to March 2023 at the Medical College hospital. It included 67 consecutive patients with the first episode of acute IWMI within 24 h of symptom onset.

### Definitions

- Inferior wall myocardial infarction is defined as ischemic cardiac pain lasting more than 30 min, characteristic ST-segment elevation of 0.1 mv or more in two or more inferior lead.
- RV infarction is defined as ST-segment elevation 0.1 mv or more in V4R in ECG taken within 6 h of symptoms onset. Significant proximal RCA stenosis is defined in coronary angiogram by the presence of occlusion 70% stenosis or more, dissected plaque, or acute thrombosis in RCA.

#### Inclusion Criteria

- Patients with the first episode of acute IWMI onset of symptoms within 24 h and admitted to SMS hospital.

#### Exclusion criteria

- All thrombolysed and previous PCI /CABG patients. Previously documented-
- Abnormal ventricular function
- Left bundle branch block
- Atrial fibrillation
- Valvular heart disease is more than mild.
- Pulmonary hypertension (all primary and secondary etiology)

### All patients underwent

Full history taking, Electrocardiogram(left and right side ECG), CK-MB (Creatine kinase-muscle brain) troponin I, and echocardiographic assessment of RV function were done within 24 h of onset of symptoms. Echocardiographic measurement was performed according to guidelines of the American Society of Echocardiography for assessment of RV function the following parameters were measured: TAPSE, tissue Doppler velocities from RV free wall, and MPI.

#### 4.1 TAPSE

In the apical 4-chamber view, the M-mode cursor was placed through the tricuspid annulus at the RV free wall. From M-mode tracing the longitudinal motion of the annulus at peak systole was measured.

**Figure 1.**
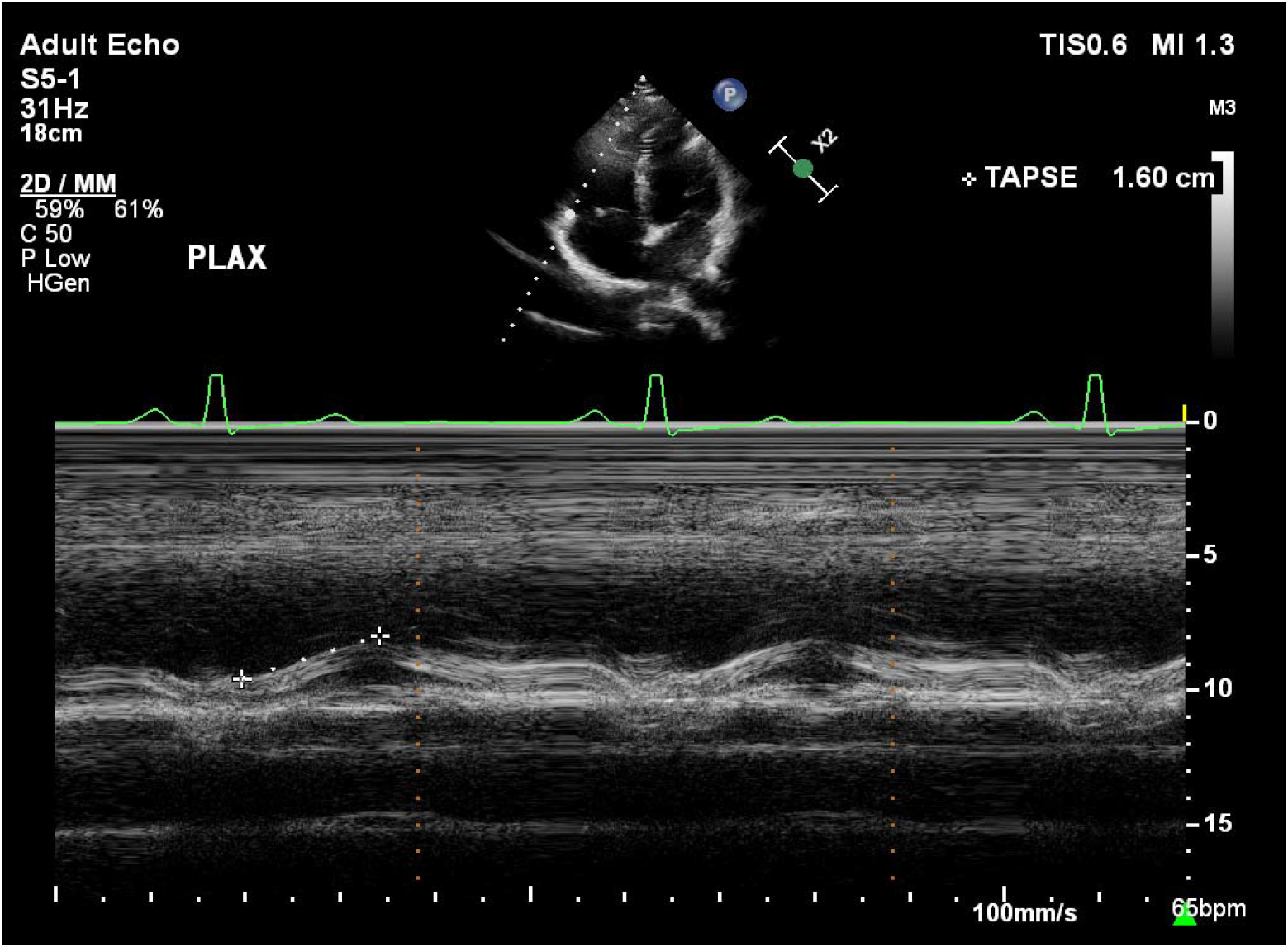
Measurement of TAPSE in the apical 4-chamber view, with M-mode.

#### 4.2 MPI by pulsed-wave Doppler method (MPI-PW)

In the apical 4-chamber view, pulsed wave Doppler tricuspid flow velocities are recorded by placing the sample volume between the leaflet tips in the center of the flow stream. Doppler beam aligned parallel to RV inflow and measurements were taken at end expiration. Tricuspid early rapid filling velocity (E), peak atrial contraction velocity (A), and tricuspid valve closure opening time (TCO) were measured as the time interval from tricuspid valve closure marked at the end of A wave to tricuspid valve opening marked at the beginning of E wave in the next cardiac cycle in pulsed wave doppler tracing. A pulsed wave doppler of the RV outflow tract was taken by placing the sample volume in the outflow tract.

Ejection time (ET) was calculated as the time from onset to cessation of flow. MPI was calculated as TCO-ET divided by ET. Beats with less than 5% variation in R–R interval were taken to allow accurate measurement of MPI. Pulsed wave tissue Doppler was acquired by placing a TDI cursor on the RV free wall at the level of the tricuspid annulus. Tissue Doppler Systolic annular velocity (S’) was measured with the movement of the annulus toward the apex during systole.

In the movement of the annulus toward the base during diastole, two major negative waves were measured: one during early diastole (E’) and one during late diastole (A’). Ejection time (ET)-duration of (S’) wave was measured. Isovolumic relaxation time (IRT)- Time between the end of (S’) and the beginning of (E’). Isovolumic contraction time (ICT) - Time between the end of (A’) and the beginning of (S’). Right ventricular MPI is calculated as (IRT+ ICT)/ET.

#### 4.3 Coronary angiogram

Within one month patients underwent coronary angiogram and were divided into two groups based on angiographic findings, group A and group B with significant proximal RCA stenosis & without significant proximal RCA stenosis respectively. Follow up for 1 month whether the patient underwent intervention or not.

### Statistical analysis

Independent-samples t-test of significance was used when comparing two means and the Chi-square (X2) test of significance was used to compare proportions between two qualitative parameters. p values < 0.05 was considered significant.

## RESULTS

In our study, the total participants were 67 among them 25 were with proximal RCA stenosis and 42 were without proximal RCA stenosis.

The mean age of patients with proximal RCA stenosis was 64.2±8.6 years and without proximal RCA stenosis was 59.8±9.8 years. In our study, RV involvement was found to be statistically significant high among the patients with proximal RCA stenosis while other sociodemographic profiles and characteristics were not found to be statistically significant different between both groups. (figure 2, Table 1)

**Table 1:** Sociodemographic and patients characteristics:

|  | With prox. RCA stenosis<br>(n=25) |  | Without prox. RCA stenosis<br>(n=42) |  | p-value |
| --- | --- | --- | --- | --- | --- |
|  | n | % | N | % |  |
| Age<br>(Mean±SD) | 64.2±8.6 |  | 59.8±9.8 |  | 0.069 |
| Gender |  |  |  |  |  |
| Male | 16 | 64.0% | 29 | 69.0% | 0.670 |
| Female | 9 | 36.0% | 13 | 31.0% |  |
| RV involvement | 23 | 92.0% | 6 | 14.3% | 0.001 |
| Smoking history | 19 | 76.0% | 33 | 78.6% | 0.807 |
| Hypertension | 9 | 36.0% | 18 | 42.9% | 0.580 |
| DM | 8 | 32.0% | 11 | 26.2% | 0.610 |
| Dyslipidaemia | 13 | 52.0% | 21 | 50.0% | 0.874 |

**Figure 2:**
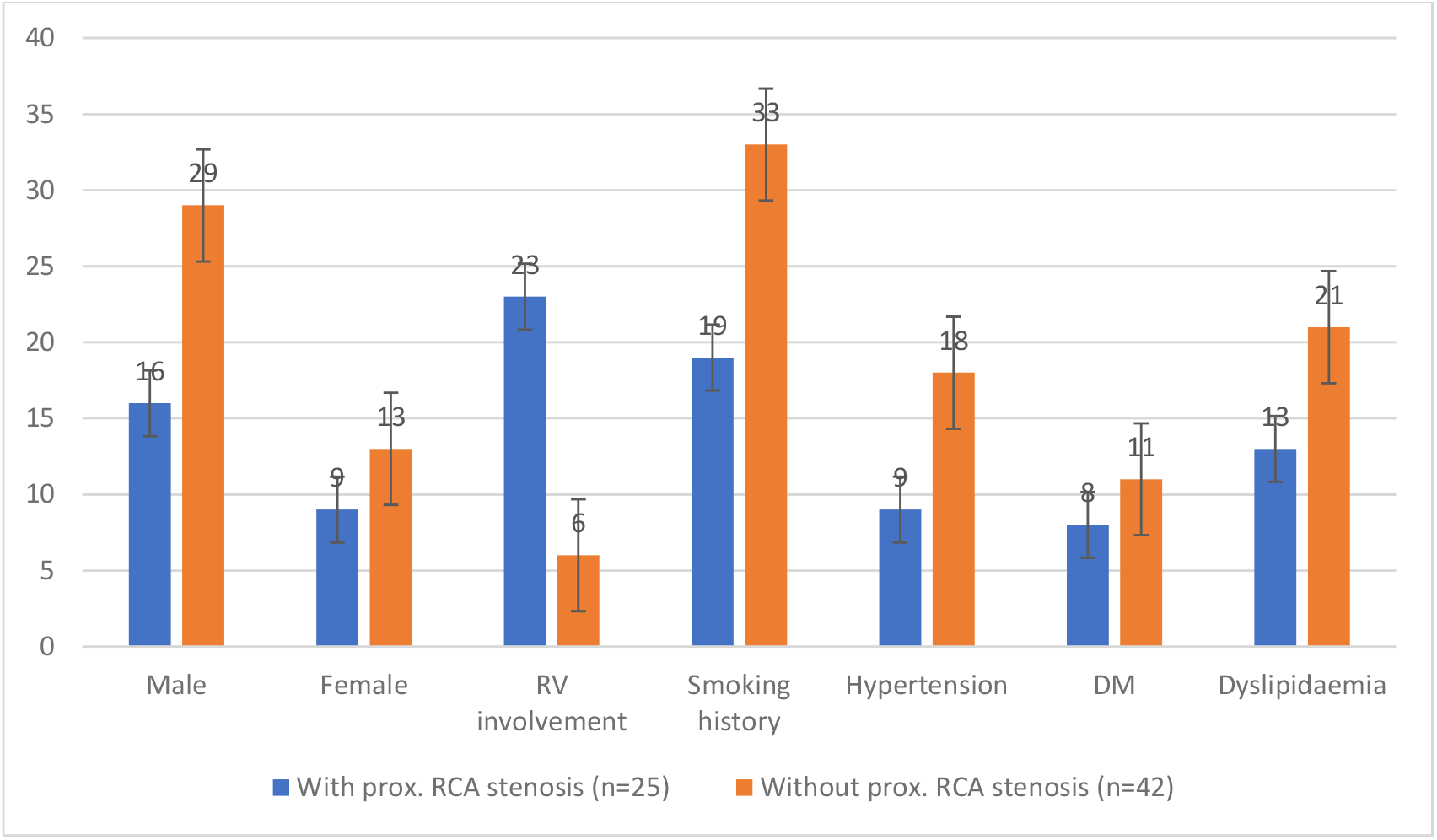
Sociodemographic and patients characteristics:

Our study found that a statistically significant high SBP, DBP, and HR were found among the patients without proximal RCA stenosis while a statistically significant more patient with raised JVP, ST elevation V4R, RV diastolic dysfunction, arrhythmias, and patients who underwent intervention were found among the group with proximal RCA stenosis. (Table 2) NT-proBNP, and Troponin I were found to be statistically significant high among the participants with proximal RCA stenosis. (Table 3) Patients with proximal RCA stenosis had a statistically significant low TAPSE-M, ET-PW, ET by TDI, S, E, and RVFAC while a statistically significant high TCO, MPI by PW, IRT by TDI, ICT by TDI, and MPI by TDI on ECHO compare to patients without RCA stenosis. (Table 4)

**Table 2:**
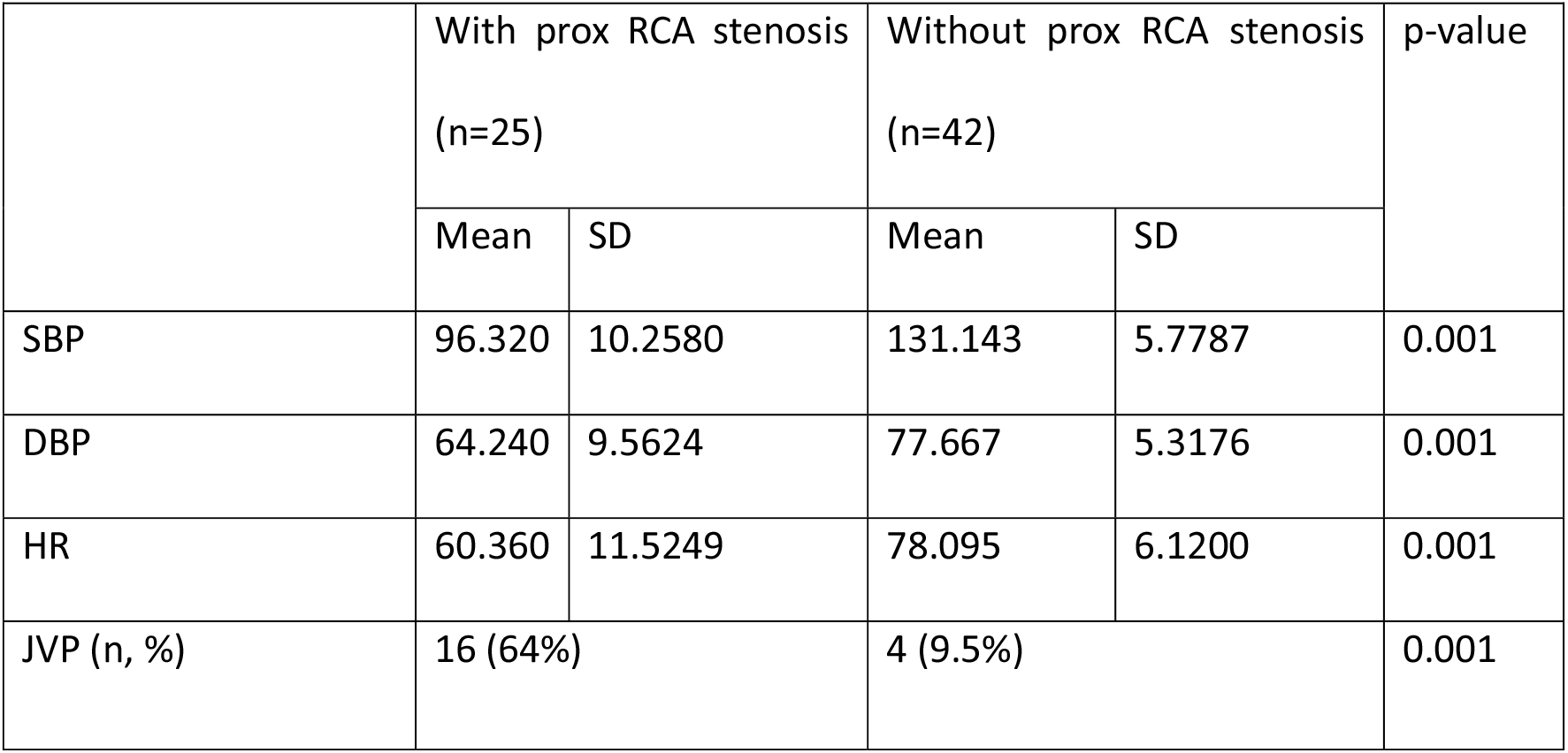
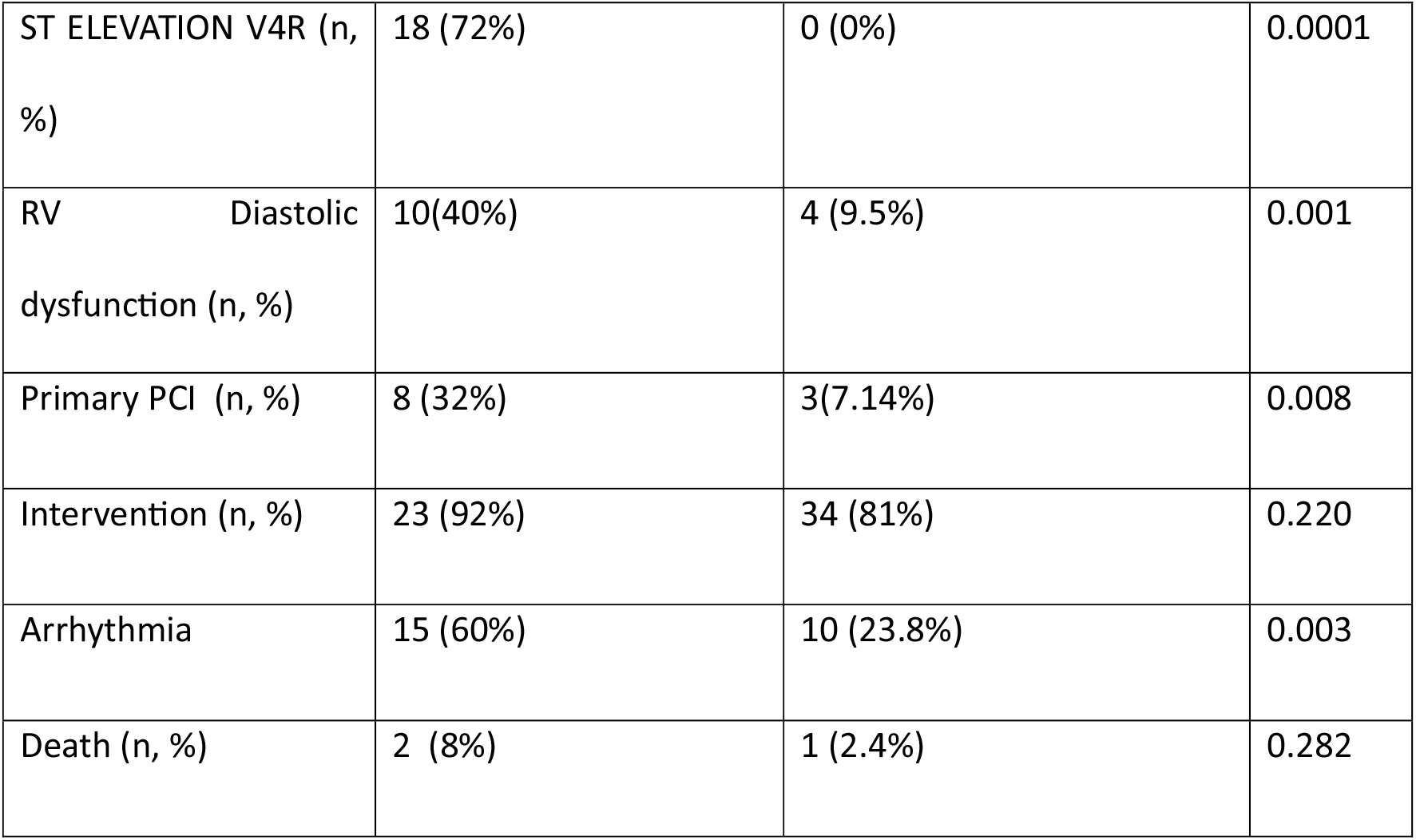
Comparison of vitals and clinical parameters.

|  | With prox RCA stenosis<br>(n=25) |  | Without prox RCA stenosis<br>(n=42) |  | p-value |
| --- | --- | --- | --- | --- | --- |
|  | Mean | SD | Mean | SD |  |
| SBP | 96.320 | 10.2580 | 131.143 | 5.7787 | 0.001 |
| DBP | 64.240 | 9.5624 | 77.667 | 5.3176 | 0.001 |
| HR | 60.360 | 11.5249 | 78.095 | 6.1200 | 0.001 |
| JVP (n, %) | 16 (64%) |  | 4 (9.5%) |  | 0.001 |

Table 2: Comparison of vitals and clinical parameters
|  |  |  |  |
| --- | --- | --- | --- |
| ST ELEVATION V4R (n, %) | 18 (72%) | 0 (0%) | 0.0001 |
| RV Diastolic dysfunction (n, %) | 10(40%) | 4 (9.5%) | 0.001 |
| Primary PCI (n, %) | 8 (32%) | 3(7.14%) | 0.008 |
| Intervention (n, %) | 23 (92%) | 34 (81%) | 0.220 |
| Arrhythmia | 15 (60%) | 10 (23.8%) | 0.003 |
| Death (n, %) | 2 (8%) | 1 (2.4%) | 0.282 |

**Table 3:** Comparison of biomarkers.

|  | With prox RCA stenosis<br>(n=25) |  | Without prox RCA stenosis<br>(n=42) |  | p-value |
| --- | --- | --- | --- | --- | --- |
|  | Mean | SD | Mean | SD |  |
| NT-proBNP (ng/L) | 579.36 | 158.838 | 394.428 | 138.291 | 0.0001 |
| Troponin I (ug/L) | 6.448 | 1.904 | 5.112 | 1.649 | 0.004 |
| CK-MB (IU/L) | 52.880 | 25.823 | 45.071 | 18.979 | 0.160 |

**Table 4:**
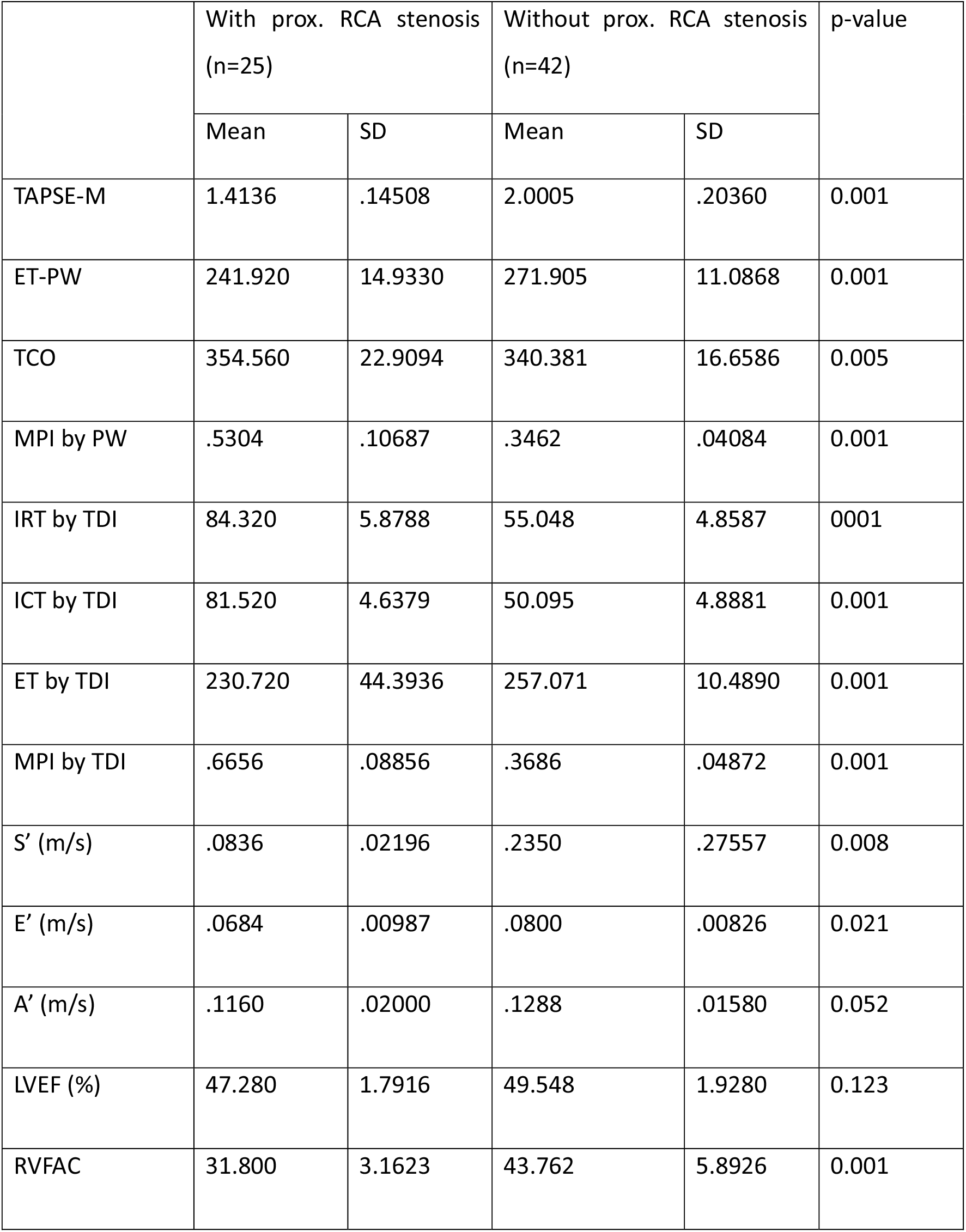
Comparison of ECHO parameters.

In the present study, maximum sensitivity to detect RCA stenosis was found by TAPSE-M mode (92%) followed by MPI by TDI (88%), and maximum 100% specificity to detect RCA stenosis was found by S’ by TDI < 10 s, MPI by TDI and ST elevation V4R.(Table 4, Figure 3,4)

**Figure 3:**
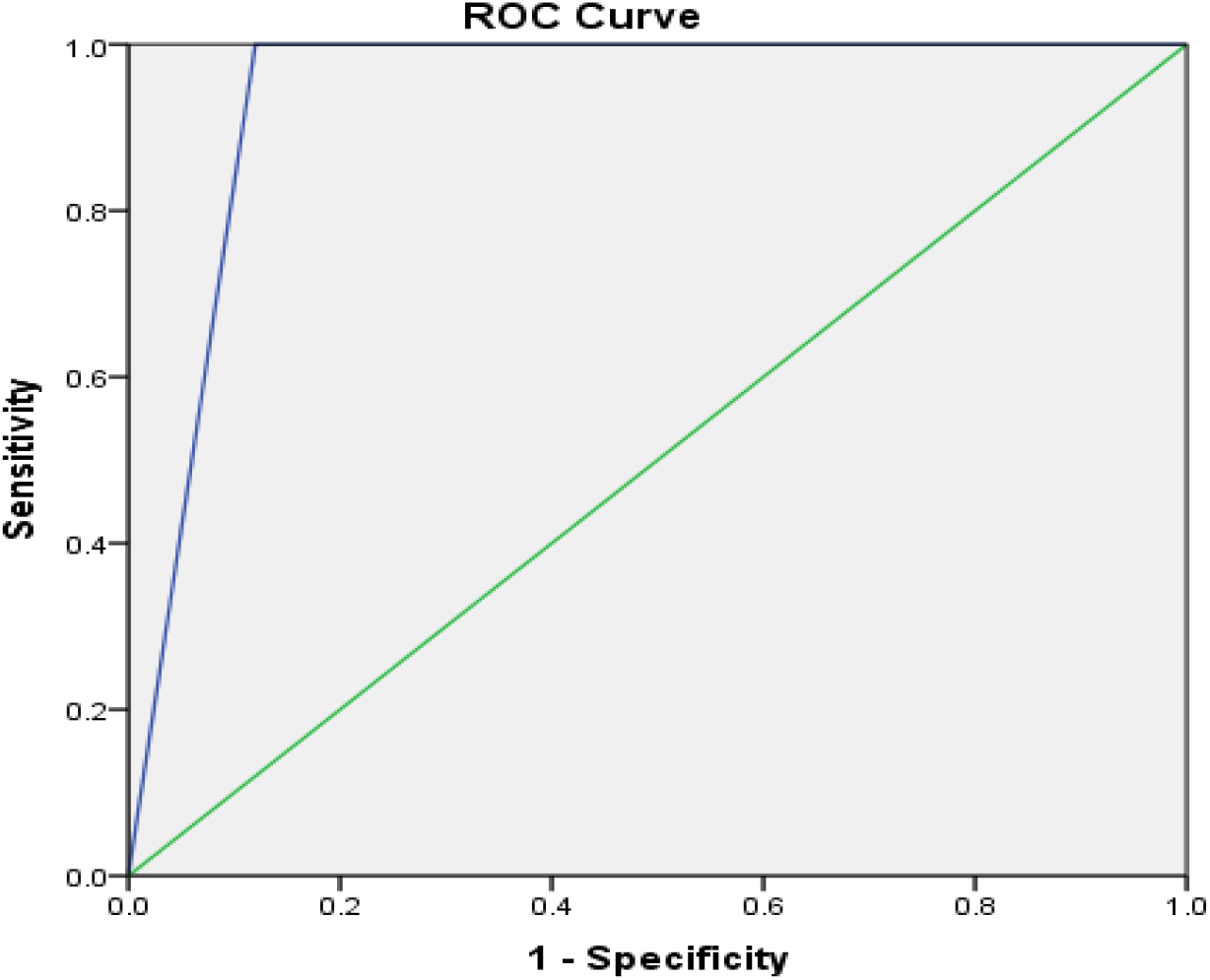
ROC for MPI-TDI.

**Figure 4:**
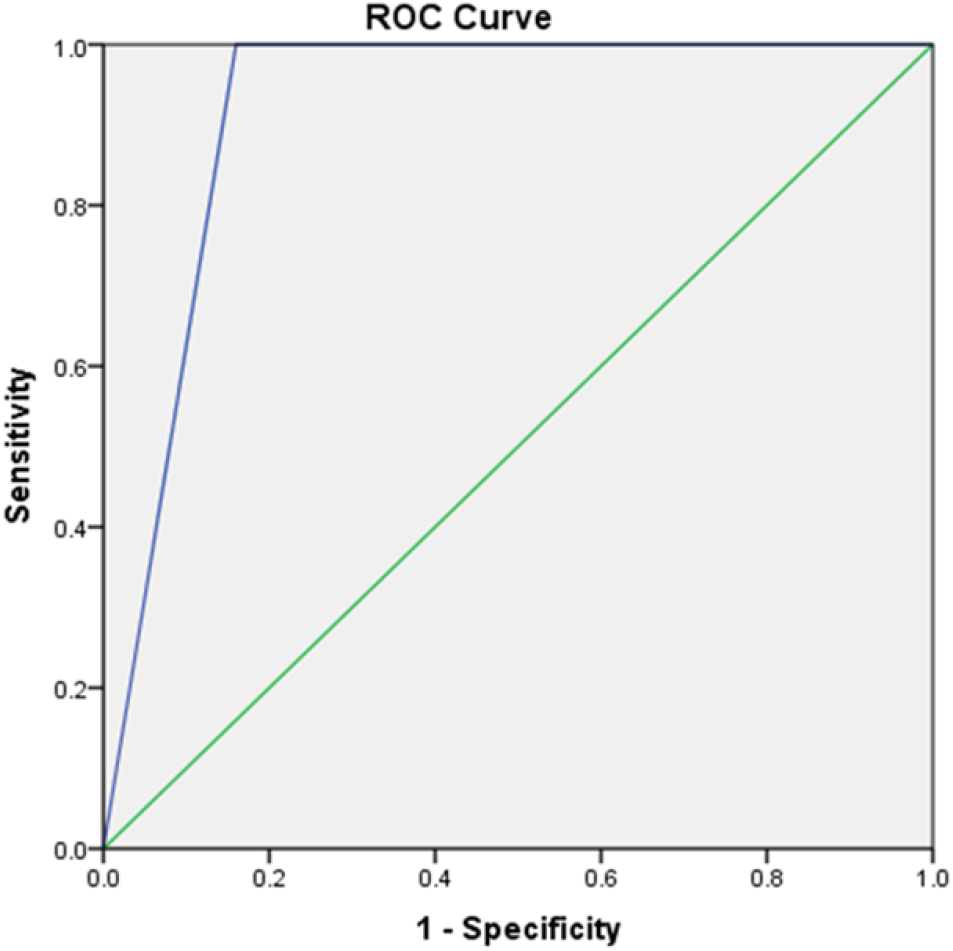
ROC for S’ <10 cm/s.

**Table 5:** Accuracy of ECHO parameters to detect proximal RCA stenosis.

| Parameter | Sensitivity | Specificity | PPV | NPV | p-value |
| --- | --- | --- | --- | --- | --- |
| $S'$ by TDI < 10 s | 84% | 100% | 100% | 91.3% | 0.0001 |
| TAPSE M mode<br>(Abnormal <1.7cm) | 92.0% | 90.5% | 85.2% | 95.0% | 0.0001 |
| MPI by PW | 84% | 92.9% | 87.5% | 90.7% | 0.0001 |
| MPI by TDI | 88% | 100% | 100% | 93.3% | 0.0001 |
| Raised JVP | 80% | 90.5% | 80% | 80.9% | 0.0001 |
| ST elevation V4R | 72% | 100% | 100% | 85.7% | 0.0001 |

## Discussion

The percentage of inferior myocardial infarctions (IMI) in all acute myocardial infarctions (AMI) is thought to be between 40 to 50 percent. It is estimated that right ventricular infarction (RVI) accounts for 30% of all acute inferior myocardial infarctions of the left ventricle. Depending on the detection criteria, the incidence of RVMI varies. According to autopsy investigations, between 24 and 34 percent of the time, fatal inferior left ventricular infarction is followed by right ventricular infarction. [9]

RVMI may develop in more than 30% of patients with acute inferior-posterior left ventricular MI, according to non-invasive studies. Hemodynamic pattern with anatomic evidence of RVMI is more frequent than anticipated.

Out of the 67 patients with acute inferior wall MI in the current investigation, 25 (37.3%) had proximal RCA stenosis, which was consistent with Shetaya AHB et al’s findings that 40% of patients had proximal RCA stenosis. [10]

According to our study, there was no statistically significant difference in the baseline characteristics between the two groups. However, among the individuals without proximal RCA stenosis, elevated SBP, DBP, and HR were found to be statistically significant. This agrees with the findings of Shetaya AHB et al.[10] Although RV infarction does not always result in hemodynamic impairment, Witt et al.’s research showed that it is typically associated with decreased RV function [11].

In the current study, the group of patients with proximal RCA stenosis contained statistically significantly more patients with elevated JVP, ST elevation V4R, RV diastolic dysfunction, arrhythmia, and patients who had undergone intervention. Raised JVP was reported to have a sensitivity and specificity of 80% and 90%, respectively. Congested neck veins and an ST-segment elevation of 1 mm in lead V4R were statistically significant in the prediction of the proximal RCA as the offending lesion, according to Shetaya AHB et al., who corroborated the findings. To identify the proximal RCA stenosis in this investigation, the specificity was 95.24% and the sensitivity was 64%. The findings of Mittal et al., who indicated that elevated jugular venous pressure had excellent specificity (96.8%) but low sensitivity (39%) in identifying RV infarction, were in agreement with these findings [12].

Troponin I and NT-proBNP levels were significantly higher in patients with RV dysfunction compared to IWMI alone, CK-MB levels were also increased but did not reach statistical significance. Comparable prognostic values of Troponin I and NT-proBNP have been demonstrated in MI and unstable angina pectoris in previous studies by Katlandur H et al. and Lindahl B et al. [13, 14]

When the ECHO parameters were evaluated in these patients, it was discovered that patients with proximal RCA stenosis had significantly lower values for TAPSE-M, ET-PW, ET by TDI, S, E, and RVFAC than patients without proximal RCA stenosis, while significantly higher values were found for TCO, MPI by PW, IRT by TDI, ICT by TDI, and MPI by TDI.

Patients with proximal RCA stenosis exhibited statistically significant low TAPSE-M and sensitivity, the specificity of TAPSE to detect the proximal RCA stenosis was 92% and 90.5%, respectively, when the ECHO parameters were examined in these patients. The Shetaya AHB et al. [10] TAPSE was significantly lower in patients with proximal RCA lesions, which supports this. Although there were fewer patients and no angiographic association, Alam M et al demonstrated a strong correlation between TAPSE and ECG evidence of RV infarction [15]. In inferior wall MI, TAPSE was likewise a reliable predictor of mortality [16]. TAPSE and right ventricular ejection fraction from radio-nuclides have a strong association, according to Kaul et al. [17]

When compared to patients without proximal RCA stenosis, patients in our study had statistically higher TCO, MPI by PW, IRT by TDI, ICT by TDI, and MPI by TDI on ECHO. MPI’s sensitivity and specificity by TDI were 88% and 100%, respectively, while by PW they were 84% and 92.9%. Similar kinds of findings were made by Shetaya AHB et al. [10] They discovered that MPI-PW correlates with a proximal RCA lesion being considerably greater in the group with proximal RCA stenosis and that it exhibits 85.71% sensitivity and 90.48% specificity. The proximal RCA lesion and MPI-TDI also displayed a highly significant connection in this investigation. According to research by Alam et al., RV infarction in relation to inferior MI can be evaluated by tricuspid annular systolic velocity by TDI. When doing dobutamine stress echocardiography, Rambaldi et al. demonstrated that the measurement of systolic velocity obtained by TDI from the RV free wall near the lateral tricuspid annulus in the apical four-chamber view can be used to detect substantial RCA disease [18].

In the present investigation, the group of patients with proximal RCA stenosis contained statistically significantly more patients with ST elevation V4R. ST elevation in V4R was reported to have a sensitivity and specificity of 72% and 100%, respectively. Numerous investigations, including those by Antman et al., who claimed that right-sided ST-segment elevation, especially in lead V4R, closely correlates with blockage of the proximal RCA and is symptomatic of acute RV damage [19], supported this. According to Zehender et al., ST-segment elevation in V4R exhibited a sensitivity and specificity of 88% and 78%, respectively, for concomitant RV infarction [20]. ST-segment elevation in lead III more than lead II is 97% sensitive but only 70% specific for right ventricular infarction, according to Saw et al. [21]

Individuals with proximal RCA stenosis in the current study had a statistically significant lower S’, E’ on ECHO than individuals without RCA stenosis. It was discovered that the parameters were 84% and 100%, respectively. The Shetaya AHB et al and S’ findings that the group with proximal RCA stenosis had considerably lower S’ values confirm this. Additionally, RV-MI patients showed considerably lower peak systolic tricuspid annular systolic velocity (S’), according to Alam et al. According to Ozdemir et al. [22]

Patients with proximal RCA lesions were shown to have considerably lower S’ values than those with distal RCA or LCX lesions. With sensitivity, specificity, positive predictive value(PPV), and negative predictive value(NPV) of 63%, 88%, 81%, and 74% respectively, they chose a value of 12 cm/s as the cutoff value. According to Meluzin et al., an S’ velocity of less than 11.5 cm/s was 90% sensitive and 85% specific in predicting RV dysfunction. [23]

## Limitation

Our study had a few limitations; the most important was the small sample size. The study duration was relatively short.

## Conclusion

In patients with an ST elevation inferior MI, echocardiographic assessment of various parameters of RV functions showed significant differences between groups with or without proximal RCA stenosis. TAPSE, Tissue Doppler systolic annular velocity, and myocardial performance index are easy to perform, sensitive markers of RV involvement, and useful in predicting proximal RCA as infarct-related artery. There was a considerable correlation between cardiac marker levels and ventricular dysfunction. It was found that high NT-proBNP and troponin I levels within the first 12 hours in patients presenting with acute IWMI might indicate associated RVMI. Patients with RV involvement in inferior wall MI were associated with more complications. Early identification of proximal RCA involvement and intervention prevents mortality and morbidity.

## Data Availability

The data used to support the findings of this study are included within the article.

